# Determinants of Condom Use Among Sexually Active Men in a High-HIV-Burden Municipality in Ghana

**DOI:** 10.64898/2026.11.12.26367946

**Authors:** Philip Yamoah Appiah, Ruth Barkey Tettey, Carmen Ortega-Santos, James Shine, Stephen Manortey

**Affiliations:** Department of Biostatistics and Bioinformatics, Milken Institute School of Public Health, The George Washington University, Washington, District of Columbia, United States of America; Department of Exercise and Nutrition Sciences, Milken Institute School of Public Health, The George Washington University, Washington, District of Columbia, United States of America; Department of Community Health, Ensign Global University, Kpong, Ghana; Oyoko Health Center, Ghana Health Service, Koforidua, Ghana

**Author notes:** Corresponding author (PA).

## Abstract

Consistent and correct condom use remains one of the most effective strategies for preventing sexually transmitted infections, including HIV, yet the determinants of male condom use are poorly documented at the district level in Ghana. This mixed-methods, cross-sectional study examined individual, sociocultural, and environmental determinants of condom use among 314 sexually active men aged 15 to 59 years, surveyed in 2016 in the Lower Manya Krobo Municipality, a district with historically elevated HIV prevalence (11.6% in 2013, 5.56% in 2019, and 4.65% in 2024). Data were collected via a structured questionnaire and focus group discussions with eight participants. In the final multivariable logistic regression model, having children was the only factor independently associated with ever having used a condom (adjusted odds ratio 3.28, 95% confidence interval 1.74 to 6.19, p<0.001); age group, marital status, community of residence, and religion are not reported as adjusted estimates because their values were unstable in the presence of sparse cells. In the focus group discussions, younger men described pregnancy prevention rather than disease avoidance as their main motivation for condom use, while religious belief and social stigma (55.4% agreeing condom use is immoral, 58.3% citing partner-suspicion stigma) emerged as persistent barriers to acceptability. A gap between high awareness of condom effectiveness (77.1%) and frequent self-reported slippage or breakage (65.2% of ever-users) suggested a skills rather than a knowledge deficit. Although the data are a decade old, the normative and religious barriers recur in more recent national surveys, and the district remains comparatively high-burden, suggesting continued programmatic relevance. HIV and STI prevention programming for Ghanaian men may be more effective when framed within a broader sexual and reproductive health and family-planning context. Engaging religious and community leaders directly and incorporating hands-on condom-use skills training rather than risk-based knowledge messaging alone may be effective.

## Introduction

Condom use remains one of the most effective and accessible strategies for preventing HIV and other sexually transmitted infections (STIs), particularly in sub-Saharan Africa, where heterosexual transmission continues to drive the epidemic [1]. In Ghana, the most recent national estimates from the Ghana AIDS Commission put adult HIV prevalence at 1.49%, with approximately 334,721 people living with HIV and 15,290 new infections recorded in 2024 alone [2,3]. Despite two decades of prevention programming, men remain markedly underserved by HIV services. A 2024 analysis of the 2022 Ghana Demographic and Health Survey (GDHS) [4] found that only around one in four Ghanaian men had ever tested for HIV and received their result, even where testing was offered free of charge [5]. Understanding what drives and what blocks condom use among men is therefore still a live public health question rather than a historical one.

A growing body of national-level evidence illustrates why this question remains open. Analysis of the 2022 GDHS found that most Ghanaian men (63%) reject the idea that contraception is solely a woman’s concern, yet a majority (59%) still believe that a woman who uses contraception may become promiscuous [4,6], an apparent tension between wanting shared responsibility and stigmatising the women who exercise it. Among Ghanaian youth, consistent condom use remains low, at around 11% in one national analysis [7]. Men who profess traditional African religion, a minority faith category distinct from Christianity or Islam, are significantly less likely to use condoms consistently than men of other faiths [7]. Exposure to family planning messaging, by contrast, is associated with a 51% higher likelihood of consistent condom use among never-married Ghanaian men, suggesting that how prevention messages are framed matters as much as whether they reach their audience at all [8].

This national picture leaves a district-level gap. In our reading of the literature, sub-national mixed-methods evidence on heterosexual condom-use determinants remains uncommon across sub-Saharan Africa, and Lower Manya Krobo Municipality, in Ghana’s Eastern Region, illustrates the gap directly. The district recorded an HIV prevalence of 11.6% at its urban sentinel site in 2013, among the highest in the country at the time [9]. More recent Ghana AIDS Commission evidence places district-level prevalence at 5.56% in 2019 and 4.65% in 2024. The district has the highest prevalence in the region [10]. Though there is a substantial decline, it is still comparatively elevated relative to the current national average of 1.49% [3], but there is little published, district-level evidence on the determinants of male condom use specifically. To the authors’ knowledge, this mixed-methods study, which combines a structured survey with focus group discussions among sexually active men in Lower Manya Krobo, is the only study to combine structured survey and qualitative data on male condom-use determinants specifically in this municipality.

Though the data analysed here were collected in 2016, it is important for current knowledge because the normative and religious barriers identified in this study recur in the national analyses reviewed above, suggesting they have not been resolved in the intervening years.

Second, while local HIV prevalence has declined since 2013, the district remains comparatively high-burden (4.65%) as of the most recent available estimate. These data therefore do not serve as a historical record but as a granular, community-level baseline for comparing subsequent national-level trends.

This study was guided by a conceptual framework integrating the Theory of Planned Behaviour [11] with an ecological view of condom-use determinants. It distinguishes individual factors, knowledge and perception factors, and environmental factors as determinants of male condom use (Fig 1). This integrated approach builds on meta-analytic evidence that attitudes and subjective norms predict condom-use intentions and that intentions in turn predict condom use, while perceived behavioural control adds little beyond intention [12]. That pattern motivated this study’s mixed-methods design: structured items to quantify individual and knowledge-related associations and focus group discussions to surface the environmental and normative context.

**Fig 1.**
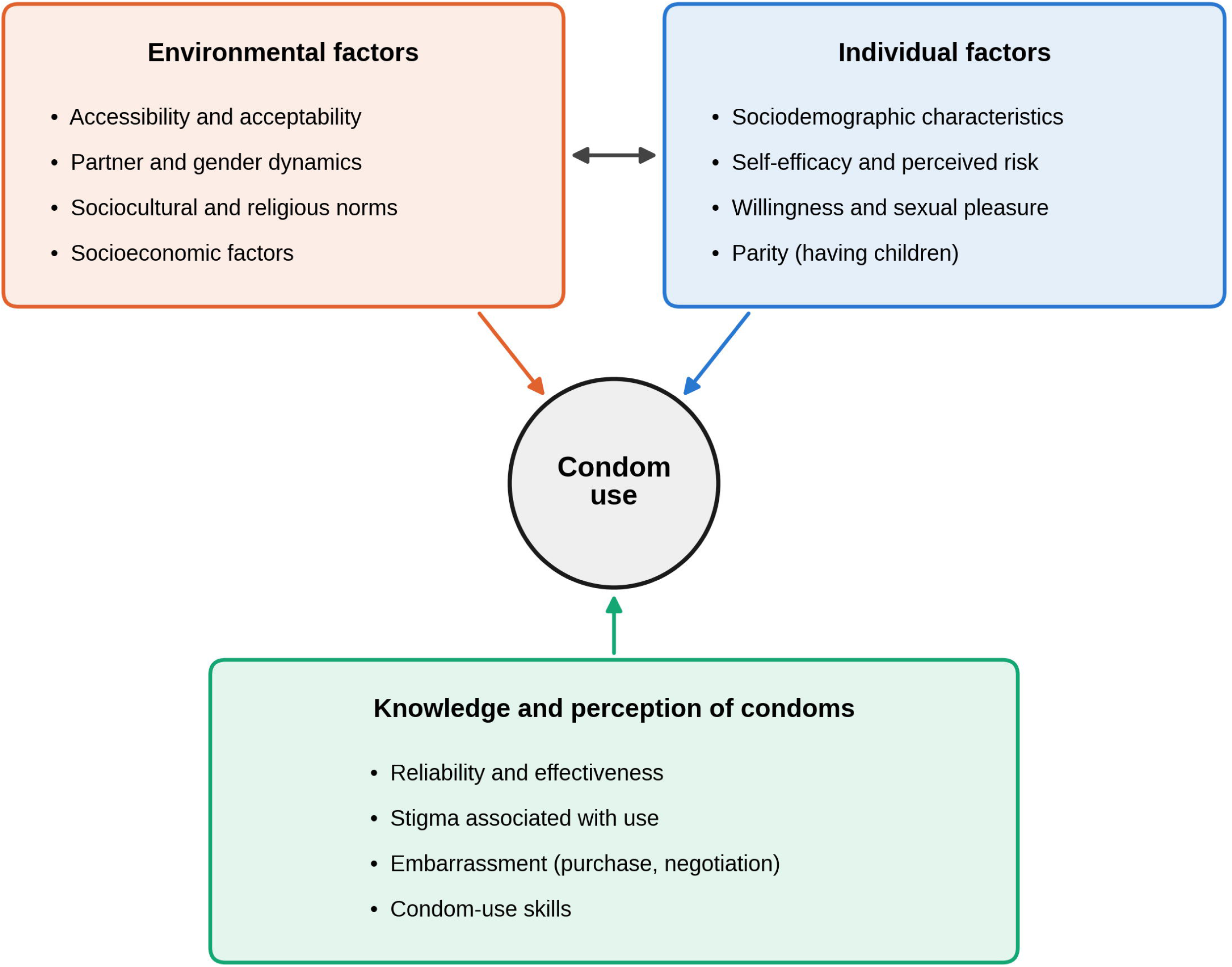
Conceptual framework of individual, environmental, and knowledge/perception factors influencing condom use.

## Materials and methods

### Study design and setting

This study examined the individual, knowledge-related, and environmental factors associated with condom use among sexually active men aged 15 to 59 years in the Lower Manya Krobo Municipality, using a mixed-methods cross-sectional design combining a structured questionnaire survey with focus group discussions.

This mixed-methods cross-sectional study conducted among sexually active men in the Lower Manya Krobo Municipality, Eastern Region of Ghana, a district with historically elevated HIV prevalence [9,10]. Quantitative data were collected via structured questionnaire interviews, and qualitative data were collected via focus group discussions (FGDs) between 11 February and 2 March 2016.

### Study population and eligibility

Eligible participants were sexually active men aged 15 to 59 years who had lived, schooled, or worked in the study area for at least one year and were capable of providing independent or parental informed consent. Researchers purposively selected six communities (Kpong, Agormanya, Akuse, and Odumase Krobo, classified as urban; Asitey and Oborpa, classified as rural) based on their population size and proximity. Within each community, men were approached at public gathering points, including commercial lorry stations, schools, workshops, drinking spots, and football parks, using a systematic convenience approach in which approximately every second eligible man encountered was invited to participate. This recruitment approach relied on purposive community selection and convenience sampling at public venues rather than household-based probability sampling.

### Sample size

The target sample size (n=355) was calculated using an estimated national male condom-use prevalence of 8%, a 95% confidence level, and a 5% margin of error, against a 2010 census population of 23,115 males aged 15 to 59 in the municipality [13]. The standard sample-size formula for an infinite or large population (n = Z²p(1−p)/e²) applied to these inputs yields approximately n = 355. This calculation in any case describes the formal target-size determination rather than the achieved precision of a probability sample, given the non-random recruitment method actually used. A total of 314 men were interviewed, 88.5% of the target.

### Data collection

Two trained research assistants conducted face-to-face interviews using a structured, closed-ended questionnaire, piloted and revised prior to fielding. The questionnaire captured sociodemographic characteristics, condom-use history, and Likert-scale items assessing knowledge of and attitudes towards condoms, adapted in part from established multidimensional condom-attitude scales [14]. Reliability was assessed via Cronbach’s alpha, with a threshold of 0.6 or above. Data were double-entered, verified, and cleaned in Microsoft Access.

2 FGDs, with four participants each, were conducted in the six communities with men aged 18 to 45 years. They were stratified by age and employment status into a younger-unemployed group and older-employed group. The discussions used a semi-structured topic guide based on preliminary quantitative findings. Each discussion was led by a moderator, supported by a note-taker who recorded observations on non-verbal cues and group dynamics. Before the discussion began, the moderator and participants agreed on ground rules regarding confidentiality, consent to audio recording, and mutual respect. Discussions were conducted in the language spoken by all participants present and lasted between one and two hours. Audio-recorded were subsequently transcribed, translated into English, and coded thematically in NVivo[15].

### Statistical and qualitative analysis

Using STATA [22], descriptive analysis on sociodemographic and behavioural characteristics was assessed. Univariate and multivariable logistic regression, with stepwise selection, examined associations between condom use (ever used) and candidate covariates. Statistical significance was interpreted at p<0.05. Univariate results for the sociodemographic covariates, with standard errors on the log-odds scale, are reported in S1 Table. Univariate p-values are unadjusted for multiple comparisons.

Four categorical covariates examined at the univariate stage (age group, marital status, community of residence, and religion) were assessed for the stability of their estimates when entered into preliminary multivariable models. Stability was judged by the width of the confidence intervals around adjusted estimates.

At the univariate stage, ethnicity was examined using the Ada group as the reference category. Categories with too few respondents to be estimable were identified and distinguished from contrasts among the larger groups. Ethnicity was eligible for the final model through the stepwise selection procedure on the same basis as the other covariates, so that omission due to ordinary non-significance could be distinguished from exclusion due to sparse-data instability. Where the multivariable model returned a single odds ratio for a variable while the univariate analysis returned a separate estimate for each category (S1 Table), the variable was taken to have entered the model as a single ordinal term rather than as a set of categorical contrasts.

Qualitative data were analysed thematically, with a coding framework derived inductively from FGD transcripts.

### Measures

The primary outcome was self-reported lifetime condom use, assessed with the question "Have you ever used a condom during sexual intercourse?" and coded as ever used (1) versus never used (0).

Individual factors comprised age, marital status, educational attainment, occupation, ethnicity, religion, community of residence, and having children (yes/no).

Knowledge and perception factors were assessed using multi-item Likert-scale subscales covering knowledge of condom reliability and effectiveness, sexual pleasure associated with condom use, stigma associated with proposing or using condoms, embarrassment about negotiating condom use, and embarrassment about purchasing condoms. Items were rated on a 5-point scale from strongly disagree to strongly agree and, for descriptive reporting, dichotomised as yes versus no. The items were adapted from the UCLA multidimensional condom attitudes scale [14].

Environmental factors comprised sociocultural, religious, and economic barriers to condom use, together with condom acquisition and storage practices and partner involvement in condom application. Among men reporting ever having used a condom, frequency of use and self-reported slippage or breakage were also recorded as indicators of condom-use skills.

### Ethics statement

This study was conducted consistent with the ethical standards of the 1964 Declaration of Helsinki and its later amendments. Ethical approval was obtained from the Ethics Review Board of Ensign Global University. Written informed consent was obtained from all participants after study aims, procedures, and confidentiality protections were explained. Participation was voluntary, and participants were free to withdraw at any time. Also, parental or guardian consent was obtained for respondents who were under 18 years of age.

## Results

### Sociodemographic characteristics

The mean age of the 314 respondents was 25.7 years (SD 6.4). The largest age group was 22 to 26 years (35.7%). Respondents were drawn from six communities, with the largest proportions from Kpong (28.0%) and Agormanya (21.0%). The majority identified as Krobo (51.9%) or Ewe (21.3%) ethnicities. Most men had completed at least basic (junior high school) education, roughly evenly split between junior high school and senior high school, technical, or vocational attainment (32.2% each), and 2.5% had no formal education. By occupation, just over a quarter (27.7%) were students, 15.9% were civil servants, 14.0% were drivers, and 13.7% were engaged in other forms of business. The remainder were distributed across artisanal, trading, farming, fishing, and unemployed categories (Table 1). The large majority (92.7%) identified as Christian. Most men were single (63.3%), while 15.3% were married and 17.9% were cohabiting; one respondent did not report marital status. Just over a third (36.9%) reported having at least one child (Table 1).

**Table 1.** Sociodemographic characteristics of respondents (N=314).

| Characteristic | n | % |
|---|---|---|
| **Age group (years)** |  |  |
| 15 | 2 | 0.6 |
| 16–21 | 65 | 20.7 |
| 22–26 | 112 | 35.7 |
| 27–31 | 66 | 21.0 |
| 32–36 | 39 | 12.4 |
| 37–41 | 24 | 7.6 |
| 42–48 | 6 | 1.9 |
| Mean (SD) | 25.7 (6.4) | n/a |
| **Marital status**<sup>a</sup> |  |  |
| Single | 198 | 63.3 |
| Cohabiting | 56 | 17.9 |
| Married | 48 | 15.3 |
| Divorced | 6 | 1.9 |
| Separated | 5 | 1.6 |
| **Educational level** |  |  |
| None | 8 | 2.5 |
| Primary | 53 | 16.9 |
| JHS/Middle | 101 | 32.2 |
| SHS/Technical/Vocational | 101 | 32.2 |
| Tertiary | 51 | 16.2 |
| **Ethnicity** |  |  |
| Krobo | 163 | 51.9 |
| Ewe | 67 | 21.3 |
| Ada | 35 | 11.2 |
| Akan | 32 | 10.2 |
| Ga | 11 | 3.5 |
| Gonja | 4 | 1.3 |
| Dagomba | 2 | 0.6 |
| **Occupation** |  |  |
| Student | 87 | 27.7 |
| Civil servant | 50 | 15.9 |
| Driver | 44 | 14.0 |
| Business (non-trading) | 43 | 13.7 |
| Unemployed | 32 | 10.2 |
| Peasant farmer | 20 | 6.4 |
| Trader | 18 | 5.7 |
| Artisan | 9 | 2.9 |
| Auto mechanic | 7 | 2.2 |
| Fisherman | 4 | 1.3 |
| **Religion** |  |  |
| Christian | 291 | 92.7 |
| Islamic | 10 | 3.2 |
| Traditional | 4 | 1.3 |
| Other | 9 | 2.9 |
| **Has children** |  |  |
| Yes | 116 | 36.9 |
| No | 198 | 63.1 |
| **Community** |  |  |
| Kpong (urban) | 88 | 28.0 |
| Agormanya (urban) | 66 | 21.0 |
| Odumase (urban) | 64 | 20.4 |
| Akuse (urban) | 45 | 14.3 |
| Asitey (rural) | 34 | 10.8 |
| Oborpa (rural) | 17 | 5.4 |

<sup>a</sup>Marital status totals 313 (99.7%) of the 314 respondents; one respondent did not report marital status. All other characteristics total 314.

### Condom use prevalence

Two-thirds of men (67.1%, 208 of 310 with valid data on this item) reported ever having used a condom. Among the 208 ever-users, 53.8% (112) had used a condom at first intercourse. Among ever-users answering the corresponding items, 46.6% (96 of 206) had used a condom at their most recent intercourse, and 43.3% (88 of 203) had used one in the preceding four weeks; expressed against the full sample of 310 men with valid outcome data, recent use falls to 28.4%. Of the 135 men who reported having sexual partners in the six months before the survey, over half (53.3%) reported more than one partner. Married men ranked second to single men in the proportion reporting more than one partner (21.1%).

### Factors associated with condom use

In the final multivariable logistic regression model, having children was the only factor independently and significantly associated with ever having used a condom: men with children had 3.28 times the odds of condom use compared with men without children (adjusted OR 3.28, 95% CI 1.74–6.19, p<0.001). Educational level (adjusted OR 1.11, 95% CI 0.95–1.27, p=0.164) and occupational category (adjusted OR 1.05, 95% CI 0.95–1.14, p=0.331) were retained by the stepwise procedure, but neither reached statistical significance once adjusted for the other covariates in the model (Table 2).

**Table 2.** Final multivariable logistic regression model of factors associated with ever having used a condom.

| Variable | Adjusted OR | 95% CI | p-value |
|---|---|---|---|
| Having children (ref: no children) | 3.28 | 1.74–6.19 | <0.001 |
| Educational level | 1.11 | 0.95–1.27 | 0.164 |
| Occupation | 1.05 | 0.95–1.14 | 0.331 |

### Knowledge and attitudes towards condom use

Reliability analysis of the attitude subscales showed acceptable to good internal consistency. Cronbach’s alpha was 0.63 for knowledge of condom reliability and effectiveness, 0.82 for sexual pleasure associated with condom use, 0.82 for stigma associated with proposing or using condoms, 0.66 for embarrassment about negotiating and using condoms, 0.71 for embarrassment about purchasing condoms, 0.61 for sociocultural and economic barriers, and 0.72 for frequency of condom use. Most men (77.1%) agreed that condoms are effective in protecting a partner from disease, and awareness of the female condom was similarly high (95.8%), though only 16.6% reported a partner had ever used one. Common barriers were widely endorsed, however, two of them by a majority: 58.3% agreed that a partner might suspect infidelity if a man proposed condom use, 55.4% agreed that condom use is immoral, and 44.3% agreed that condoms make a man look silly in front of a partner. Just over a third (36.9%) agreed that condoms reduce sexual pleasure (Table 3).

**Table 3.** Knowledge and attitudes towards condom use among respondents (N=314).

| Item | Agree n (%) | Disagree n (%) | Neutral n (%) |
|---|---|---|---|
| Condoms protect a partner from disease | 242 (77.1) | 55 (17.5) | 17 (5.4) |
| Condoms reduce sexual pleasure | 116 (36.9) | 166 (52.9) | 32 (10.2) |
| A partner may suspect infidelity if condom use is proposed | 183 (58.3) | 109 (34.7) | 22 (7.0) |
| Condoms make a man look silly before his partner | 139 (44.3) | 144 (45.9) | 31 (9.9) |
| Using condoms is immoral | 174 (55.4) | 123 (39.2) | 17 (5.4) |
| It is embarrassing to buy condoms in a store | 95 (30.3) | 202 (64.3) | 17 (5.4) |

### Condom use skills

Among men who had ever used a condom, 65.2% reported experiencing slippage or breakage at least once in the preceding 12 months, and 70% reported not involving their partner in applying the condom. Storage practices were often suboptimal: 28.8% typically carried condoms in a pocket, and 11.1% reported storing them in locations such as under a car seat.

### Qualitative findings

Three themes emerged from the focus group discussions (Table 4), with some variation between the younger-unemployed and older-employed discussion groups.

**Table 4.** Qualitative themes and illustrative quotations from focus group discussions.

| Theme | Description | Illustrative quotation |
|---|---|---|
| Pregnancy prevention as primary motivator | Younger men prioritised preventing pregnancy over disease prevention as the reason for condom use | "…my main concern is to prevent pregnancy because I am not much scared about the disease…" |
| Religious and moral objections | Christian religious teaching invoked as a barrier to condom acceptability | "it is immoral to me because from the onset of the generation of God, he created man and woman to have sex but he never added any material to it" |
| Peer norms and knowledge gaps | Declining peer acceptability of condoms, and persistence of misconceptions such as withdrawal as a substitute method | "the guys pull their penis out of the vagina when the sperm is about to come… so they think they are expected in that…" |

First, pregnancy prevention, not STI or HIV protection, was the primary motivation for condom use described by younger men: *"…my main concern is to prevent pregnancy because I am not much scared about the disease, and thus the mindset of most men*” (participant, focus group discussion). Multiple-partnership behaviour was described candidly by younger participants as a further driver of condom use: "*We are sexually active and we don’t date one lady; we date several ladies, and now there are sicknesses around, so we should always use a condom to protect ourselves”* (younger participant, focus group discussion), though the same group also described situational risk judgements that could override this intention, such as assessing a new partner as low-risk based on her perceived lifestyle.

Second, religious belief was described as a barrier to acceptability: "*It is immoral to me because from the onset of the generation of God, he created man and woman to have sex but he never added any material to it*” (participant, focus group discussion).

Third, participants in the younger group described declining condom acceptability among their peers and gaps in practical knowledge, including a belief in withdrawal as an adequate substitute: “*the guys pull their penis out of the vagina when the sperm is about to come… so they think they are expected in that… they don’t go in for the condom”* (younger participant, focus group discussion), alongside a lack of discreet or youth-friendly ways to purchase condoms without embarrassment.

## Discussion

Condom use remains central to HIV and STI prevention in Ghana, where the epidemic, though far from its historical peak, is still active: an estimated 15,290 Ghanaians acquired HIV in 2024 alone [3], and men remain markedly underengaged with prevention services, with only around one in four having ever tested for HIV and received their result [5]. Against this backdrop, the present study’s district-level, mixed-methods evidence on male condom-use determinants in a historically high-prevalence municipality remains substantively relevant.

To the authors’ knowledge, this is the only study to combine structured survey and qualitative data on male condom-use determinants specifically in Lower Manya Krobo. It therefore offers a granular, community-level baseline that complements, rather than duplicates, subsequent national surveys such as the 2022 GDHS.

The finding that having children was the strongest independent predictor of condom use is consistent with the qualitative theme that pregnancy prevention, not disease avoidance, was the men’s primary stated motivation. This evidence sits within a national context in which men’s attitudes towards contraception are themselves unsettled: a majority of Ghanaian men reject the idea that contraception is solely a woman’s concern, yet a similar majority still believe that a woman who uses contraception may become promiscuous [6], and consistent condom use among Ghanaian youth remains low nationally, at around 11% in one recent analysis [7]. Earlier multi-country evidence from adolescents aged 15 to 19 in Burkina Faso, Ghana, Malawi, and Uganda points the same way. Among young women who did not use a condom at last sex, wanting to become pregnant was the single most common reason given in both Burkina Faso and Ghana [16]. These findings may suggest that framing condom promotion primarily around HIV and STI risk may not align with how men themselves reason about condom use. This supports the interpretation that condom promotion may be more effective when embedded within a broader sexual and reproductive health and family-planning framework rather than presented solely as an HIV-specific intervention. And this is consistent with the evidence that exposure to family-planning messaging is associated with substantially higher rates of consistent condom use among never-married Ghanaian men [8].

Religious belief and social norms emerged, both qualitatively and in the attitude items (55.4% agreeing condom use is immoral, 58.3% citing partner-suspicion stigma), as persistent barriers to condom acceptability. This mirrors the finding of the analysis of the 2014 GDHS that Ghanaian men who profess Traditional African religion are significantly less likely to use condoms consistently than men of other faiths [7]. This sits alongside national-level evidence that apparent religious differences in Ghanaian men’s sexual risk-taking are largely explained by socioeconomic selection rather than by religion itself [17]. Evidence from rural Ethiopia also points to a related mechanism, in which married couples rarely used condoms because marriage was itself treated as a form of protection against infection [18]. Together, this suggests that religious and normative barriers to condom acceptability, though not identical in form across settings, recur widely enough to continue warranting targeted engagement.

In this sample, married men ranked second only to single men in the proportion reporting more than one partner (21.1%). That sits awkwardly beside evidence from Nigeria’s 2018 Demographic and Health Survey, where married men showed markedly lower consistent condom use than their unmarried counterparts, a gap the authors attributed to married men being more likely to be trying to conceive, to have fewer partners, and to engage in less risky sexual behaviour [19]. Perceived personal risk has separately been identified as strongly associated with condom use among adolescents in four sub-Saharan African countries, including Ghana [20], which may help explain why married men in this sample, despite being formally in a lower-risk relationship category, did not behave as a uniformly low-condom-use group. Williamson and colleagues, working with married couples in Kampala, found that husbands who accepted condoms gave a mix of protective and self-interested reasons, ranging from the welfare of a partner and children through to the maintenance of concurrent relationships [21]. This cross-country pattern underscores that marital status alone is an unreliable proxy for perceived risk and that programming should assess concurrency and perceived risk directly, rather than assuming that married men are a lower-priority population for condom promotion.

The widespread awareness of condom effectiveness (77.1%) alongside a substantial rate of self-reported slippage or breakage (65.2% of ever-users) and low partner involvement in application (only 30% sought partner assistance) is, descriptively, more consistent with a skills gap than a knowledge gap, though this comparison rests on separate descriptive percentages rather than a formally tested model. In our experience of prevention programming in this setting, that distinction is not always made explicit.

The pattern observed here suggests hands-on, skills-based condom demonstration and negotiation training may address a gap that knowledge-focused messaging alone does not reach.

Taken together, these findings carry direct programmatic implications for the Ghana Health Service and district health directorates operating in similarly high-burden municipalities. A pregnancy and family-planning-led messaging frame, hands-on skills training delivered through venues such as workplaces and community gathering points, and direct engagement with religious and community leaders are each more consistent with what this study found than risk-based knowledge messaging alone.

The relevance of these findings should be weighed against the age of the underlying data. Antenatal sentinel surveillance at the district’s urban site recorded an HIV prevalence of 11.6% in 2013 [9], while district population prevalence was estimated at 5.56% in 2019 [10]. These two figures are measured on different populations and are not directly comparable, but both place Lower Manya Krobo well above the national adult prevalence of 1.49% [3]. The normative and religious barriers identified here also recur in national survey data collected in 2022 [6] and in a recently published analysis of the 2014 GDHS [7].

### Strengths and limitations

The principal strength of this study is its mixed-methods design. The survey identified having children as the only factor independently associated with ever having used a condom, and the focus group discussions suggested a plausible explanation for this association: participants framed condom use primarily as a means of preventing pregnancy rather than infection. The qualitative data also helped explain the normative barriers the survey measured, showing how religious belief and social stigma shape the acceptability of condoms in ways that structured items could quantify but not explain. This integration of the two strands is the study’s central contribution.

Several limitations should be considered. First, the cross-sectional design precludes causal inference. All associations reported here, including that between having children and ever having used a condom, should be interpreted as associations rather than as evidence of a causal pathway. Second, participants were recruited by convenience sampling at public venues rather than through household-based probability sampling. Men who frequent such venues may differ systematically from those who do not, which may limit the generalisability of the findings to the wider male population of the municipality. Third, sexual behaviour was self-reported and is therefore subject to recall bias and social desirability bias. Given the sensitivity of the topic and the stigma attached to condom use in this setting, the direction of any social desirability bias is uncertain: some men may have over-reported condom use to appear responsible, while others may have under-reported it to avoid association with infidelity or immorality.

## Conclusion

Among sexually active men in a comparatively high-HIV-burden municipality in Ghana, having children was the only factor independently associated with ever having used a condom in the final model. In the focus group discussions, younger men described pregnancy prevention rather than disease avoidance as their main motivation for condom use, and participants identified religious belief and social stigma as persistent barriers to acceptability. Although most men recognised that condoms are effective, slippage and breakage were commonly reported, suggesting a gap in condom-use skills rather than in knowledge.

In this and similar settings, HIV and STI prevention programmes for men may be more effective if they are framed within a broader sexual and reproductive health and family-planning context, engage religious and community leaders to address normative barriers directly, and include hands-on condom-use skills training rather than relying on risk-based knowledge messaging alone.

## Data Availability

All data produced in the present work are contained in the manuscript

## Acknowledgments

These data were originally collected for the first author’s MPH thesis at Ensign Global University, Ghana, in 2016. The authors thank the study participants and the field.

## Supporting information

**S1 Table. Univariate logistic regression of factors associated with ever having used a condom (N=314).** Odds ratios, 95% confidence intervals, and p-values for all candidate covariates, with standard errors on the log-odds scale derived from the reported confidence limits.

