## Supplementary material for "Determinants of Condom Use Among Sexually Active Men in a High-HIV-Burden Municipality in Ghana": S1 Table

**S1 Table. Univariate logistic regression of sociodemographic factors associated with ever having used a condom.**

| Variable | Odds ratio | 95% CI | SE (log odds) | p-value |
|---|---|---|---|---|
| **Marital Status** | | | | |
| Co-habiting | 1.00 | reference | | |
| Divorced | 1.66 | 0.170 to 15.56 | 1.152 | 0.65 |
| Married | 2.27 | 0.790 to 6.50 | 0.538 | 0.12 |
| Separated | | not estimable (dropped; n=5) | | |
| Single | 0.46 | 0.240 to 0.91 | 0.340 | 0.02 |
| **Locality** | | | | |
| Agormanya | 1.00 | reference | | |
| Akuse | 2.77 | 1.060 to 7.12 | 0.486 | 0.03 |
| Asitey | 3.31 | 1.130 to 9.69 | 0.548 | 0.02 |
| Kpong | 1.16 | 0.590 to 2.27 | 0.344 | 0.66 |
| Oborpa | 0.31 | 0.100 to 0.94 | 0.572 | 0.04 |
| Odumase | 0.83 | 0.410 to 1.69 | 0.361 | 0.61 |
| **Educational Level** | | | | |
| JHS/Middle Form 1-4 | 1.00 | reference | | |
| None | 1.64 | 0.310 to 8.56 | 0.847 | 0.55 |
| Primary(1-6) | 0.59 | 0.290 to 1.16 | 0.354 | 0.13 |
| SHS/Tech/Voc. Sch. | 1.23 | 0.590 to 2.58 | 0.376 | 0.56 |
| Six Form/Poly/University | 2.30 | 1.000 to 5.32 | 0.426 | 0.05 |
| **Ethnic Group** | | | | |
| Ada | 1.00 | reference | | |
| Akan | 0.88 | 0.270 to 2.90 | 0.606 | 0.84 |
| Dagomba | | not estimable (dropped; n=2) | | |
| Ewe | 0.46 | 0.170 to 1.22 | 0.503 | 0.12 |
| Ga | 0.45 | 0.100 to 1.99 | 0.763 | 0.29 |
| Gonja | | not estimable (dropped; n=4) | | |
| Krobo | 0.45 | 0.180 to 1.11 | 0.464 | 0.08 |
| **Religious background** | | | | |
| Christianity | 1.00 | reference | | |
| Islamic | 0.46 | 0.130 to 1.63 | 0.645 | 0.23 |
| Traditional | 1.38 | 0.140 to 13.50 | 1.166 | 0.77 |
| **Occupation** | | | | |
| Business Personnel | 1.00 | reference | | |
| Civil Servant | 1.35 | 0.260 to 6.85 | 0.835 | 0.71 |
| Driver | 1.87 | 0.370 to 9.47 | 0.827 | 0.44 |

| Variable | Odds ratio | 95% CI | SE (log odds) | p-value |
|---|---|---|---|---|
| Fisherman | 1.19 | 0.230 to 5.99 | 0.832 | 0.83 |
| Peasant Farmer | 0.75 | 0.032 to 17.50 | 1.608 | 0.85 |
| Trader | 1.80 | 0.290 to 11.16 | 0.931 | 0.52 |
| Unemployed | 1.43 | 0.270 to 7.56 | 0.850 | 0.67 |
| Student | 1.47 | 0.300 to 7.03 | 0.805 | 0.62 |
| **Any Child** | | | | |
| No | 1.00 | reference | | |
| Yes | 2.72 | 1.580 to 4.66 | 0.276 | <0.001 |
| **Age group** | | | | |
| 15 | 1.00 | reference | | |
| 16 - 21 | 0.75 | 0.040 to 12.63 | 1.468 | 0.84 |
| 22-26 | 2.29 | 0.130 to 37.76 | 1.447 | 0.56 |
| 27 - 31 | 3.92 | 0.220 to 67.00 | 1.459 | 0.34 |
| 32 - 36 | 11.33 | 0.550 to 230.54 | 1.540 | 0.11 |
| 37 -41 | 1.18 | 0.065 to 21.70 | 1.482 | 0.91 |
| 42- 46 | 3.00 | 0.080 to 107.44 | 1.837 | 0.54 |
| 47 – 51 | | not estimable (dropped) | | |
